# EFFECT OF LINEAR GROWTH RATE AND CHANGE IN BODY MASS INDEX IN CHILDHOOD AND ADOLESCENCE ON BLOOD PRESSURE IN AFRO-CARIBBEAN YOUTH: THE VULNERABLE WINDOWS COHORT STUDY

**DOI:** 10.1101/2021.04.17.21255680

**Authors:** Trevor S. Ferguson, Tamika Y. Royal-Thomas, Lisa Chin-Harty, Minerva M. Thame, Terrence E. Forrester, Clive Osmond, Michael S. Boyne, Rainford. J. Wilks

**Affiliations:** Caribbean Institute for Health Research, The University of the West Indies, Mona, Kingston 7, Jamaica; Department of Mathematics, Faculty of Science and Technology, The University of the West Indies, Mona, Kingston 7 Jamaica; Department of Medicine, Faculty of Medical Sciences, The University of the West Indies, Mona, Kingston 7, Jamaica; Department of Child and Adolescent Health, Faculty of Medical Sciences, The University of the West Indies, Mona, Kingston 7, Jamaica; UWI Solutions for Developing Countries, The University of the West Indies, Mona, Kingston 7, Jamaica; Faculty of Medicine, University of Southampton, England

**Keywords:** postnatal growth, blood pressure, growth, Jamaica, Afro-Caribbean

## Abstract

**Background:** Faster growth velocity during childhood may increase blood pressure (BP) in adults, but there are little data from African-origin populations. We evaluated the effect of postnatal linear growth (increase in height) and change in body mass index (BMI) from birth to adolescence on systolic and diastolic blood pressure (SBP and DBP) in Afro-Caribbean youth.

**Methods:** We used data from the Vulnerable Windows Birth Cohort Study in Jamaica. Children were followed from birth, with periodic anthropometric measurements. BP measurements started at age 1-year and every six months thereafter. Analyses used BP measurements (mmHg) from age 15-21 years. Linear growth and change in BMI measurements were calculated for: early infancy (0-6 months), late infancy (6 months - 2 years), early childhood (2-8 years), and later childhood (8-15 years). Conditional analyses were used to compute growth rates (as z-scores). Linear mixed models were used to estimate the effect of growth rates on BP.

**Results:** Analyses included 365 individuals (162 males, 203 females) with mean age 16.7 years. In multivariable models, after adjustment for age, sex, birth length, gestational age and BMI at age 15 years, faster linear growth for early infancy (β=1.06, p=0.010) was significantly associated with higher SBP. For change in BMI, after adjustment for age, sex, gestational age, height and SES at birth, significant associations of higher SBP were seen for greater increase in BMI in late infancy (β=1.41, p=0.001), early childhood (β=1.22, p=0.001) and later childhood (β=0.74, p=0.035). Faster post-natal linear growth had no significant associations with DBP, but greater increase in BMI for each of the late infancy to late childhood periods was significantly associated with higher DBP. When both growth rates were modeled together, rate of change of BMI and faster linear growth in early infancy retained significance for SBP, but only change in BMI retained significance for DBP.

**Conclusion:** Faster linear growth and greater rate of increase in BMI were associated with higher SBP and DBP in Afro-Caribbean youth, but the BMI effect was stronger.

## INTRODUCTION

Growth in early life is related to blood pressure and other chronic non-communicable diseases in later life (1-7). This is supported by research conducted over the past thirty years, showing that factors in the prenatal, intra-uterine and early postnatal environment may have long term effects on human physiology which increases the risk of several non-communicable diseases, including cardiovascular disease and diabetes (1-4, 8, 9). The main mechanism may be epigenetic phenomena, which results in altered expression of DNA and thus phenotypic manifestations (3, 4, 8). For hypertension alternations in renal development, including nephron numbers, are also thought to play a role (9).

In addition to the studies looking at intra-uterine growth, several studies have suggested that growth velocity during childhood and adolescence may also influence blood pressure and other cardiovascular disease risk factors in adult life (10-18). Both faster linear growth and greater rate of gain in adiposity have been shown to influence blood pressure. The potential detrimental effect of faster linear growth has led to questions regarding the best strategy for promoting growth particularly for small for gestational age and preterm infants (10, 19-23). However, some studies have suggested that despite the potential adverse metabolic outcomes, faster catch-up growth may give short term benefits, such as fewer hospital admissions in early childhood, and benefits to social capital (attained height and education) in young adulthood (10, 22).

Most studies on the developmental origin of health and disease (DOHAD) have been conducted in developed countries. While there is an accumulating body of data from developing countries, including Jamaica (10, 24-29), data from African origin populations are still limited. In the Caribbean, studies on the early life influences on blood pressure have looked mainly on the effects of intrauterine growth and blood pressure in childhood or adolescence. To date, there are no studies evaluating the effects of postnatal growth on blood pressure in youth or adulthood in Afro-Caribbean populations. This study therefore aimed to evaluate the effect of postnatal linear growth and change in body mass index (BMI) from birth to adolescence, on systolic and diastolic blood pressure (SBP and DBP) in Jamaica.

The specific objectives were: (1) to estimate the effect of linear growth rate from birth to age 15 years on SBP and DBP among Afro-Caribbean youth in Jamaica, and (2) to estimate the effect of relative increase in BMI from birth to age 15 years on SBP and DBP among Afro-Caribbean youth in Jamaica.

## METHODS

### Data Sources and Participants

We analyzed data from the Vulnerable Windows Cohort Study (25). This cohort includes participants whose mothers were recruited during the first trimester of pregnancy and had follow up data spanning the prenatal period up to age 18-21 years. The original study, which began in 1992-1993, recruited 712 women, between the ages of 15 - 40 years, who were attending the antenatal clinic at the University Hospital of the West Indies (UHWI) and were 7-10 weeks into their pregnancy (25). Of these, 569 singleton live births were included in a follow up study and 365 had data on age, sex and blood pressure measured after age 15 years and were therefore included in this paper. The study was reviewed and approved by the Faculty of Medical Sciences / University Hospital of the West Indies Ethics Committee. Data used in this manuscript are available from the authors upon request.

### Measurements

Details of measurement protocols for the cohort have been previously published (24, 25, 30, 31). We therefore present a brief description of the measurements pertinent to the current study. Participants had detailed intrauterine measurements made by ultrasound, as well as standard anthropometric measurements taken at the time of birth. Postnatal measurements included anthropometry at six weeks, then every three months up to age 2 years, and every six months thereafter. Blood pressure was measured at age 1 year and every six months thereafter.

Weight during childhood was measured using a Weylus beam balance (CMS Weighing Equipment Ltd, London) and was recorded to the nearest 0.01 kg. Height was measured with a stadiometer (CMS Weighing Equipment Ltd) and recorded to the nearest 0.1 cm. Body mass index (BMI) was calculated as weight (kg) divided by the square of height (m). Blood pressure was measured twice using an oscillometric sphygmomanometer. The mean of the two measurements were used for the analysis.

Birth weight was measured using an electronic balance (CMS Weighing Equipment Ltd), while birth length was measured using a length board (Holtain Ltd). BMI at birth was calculated as birth weight (kg) divided by the square of birth length (m). We chose birth BMI instead of ponderal index in order to facilitate assessment of change fatness in the postnatal period. This was based on a study by Cole and colleagues which suggested that BMI may more appropriate measure of fatness in postnatal period (after 39 weeks gestation) when compared to ponderal index (32). Gestational age using the date on the last menstrual period was validated by fetal ultrasound at 14 weeks of gestation. Socioeconomic status (SES) at birth was determined using a composite score derived from a questionnaire on household possessions, crowding in the home, occupation and education level of the child’s mother and father (31). Higher scores indicated higher SES.

### Sample size and power

The available sample for this study included 365 participants with 1909 SBP measurements and 1907 DBP measurements. We obtained estimated power for the available sample using the number of SBP and DBP measurements, adjusting for clustering using the formula for design effect recommended by Lohr (*design effect = 1 + (M - 1) × ICC*, where M = number of observations per cluster and ICC = intraclass correlation coefficient (33). Calculated design effects were 1.71 for SBP and 1.75 for DBP. Adjusted sample size for SBP was 1116 and 1089 for DBP. For all calculations α = 0.05. Based on these sample sizes the estimated power for the effect of change in linear growth on SBP ranged from 18% for a regression coefficient of 0.36 to 83% with regression coefficient of 1.0. For change in BMI and SBP power estimates ranged from 25% to 99% for regression coefficient ranging from 0.44 to 1.47. Estimated power for DBP ranged from 23% to 57% with regression coefficients from 0.27 to 0.48 for change in linear growth and 9% to 80% for change in BMI with regression coefficients ranging from 0.13 to 0.63.

### Statistical Analyses

Data analysis was performed using Stata 14 (Stata Corp, College Station, Texas). Descriptive analyses yielded means and proportions of participant characteristics, and mixed effects linear regression models were used to assess associations between blood pressure and putative explanatory variables. Growth and BMI measurements were calculated for the periods 0-6 months (early infancy), 6-months to 2 years (late infancy), 2-8 years (early childhood), and 8-15 years (later childhood). Conditional analyses were used to compute relative linear growth and change in BMI within each period using methods described by Kajantie et al. (34). This involved calculating how much body size at a particular age differed from that predicted by the body size attained at an earlier age using linear regression models. The difference between the observed and the predicted values were used to represent the conditional linear growth and change in BMI for each period. These conditional ‘growth rates’ ensure that there is little correlation between ‘growth rates’ for each period and can thus be fitted in the same regression model. Pearson’s correlation coefficients for these ‘linear growth rates’ and change in BMI in each period are shown in Table S1 of the supplementary file. Values for growth in each period were standardized by converting them to z-scores. Mixed effects linear models were used to estimate the effect of linear growth and change in BMI on SBP and DBP. Assessment of sex interaction was done by including an interaction term in the initial regression models. The mixed effects models accounted for repeated measures within individuals, with random intercept (represented by the individual) and random slopes (represented by time [age] at which measurement was taken). Analyses were restricted to participants who had data on blood pressure on at least one occasion after age 15 years and data on age and sex. For other variables multiple imputation by chained equations was used to account for missing values. Models used 20 imputed data sets with estimates combined using Rubin’s rules (35). Most variables had less than 20% missing observations. Details of missing and non-missing variables are shown in Table S2 in the supplementary file.

## RESULTS

Analyses included 365 individuals (162 males, 203 females) with mean age 16.7 ± 1.2 years (range 15.0, 21.2 years). Included participants had similar birth characteristics to the other members of the cohort except for a 0.11 kg higher mean birth weight (p=0.023). Descriptive characteristics for study participants are shown in Table 1. At age 15 years, mean height was 172 cm for males and 164 cm for females. Mean BMI was 21.0 cm among males and 22.2 among females. Spaghetti plots for linear growth and BMI during childhood are shown in Figures1 and 2. In the spaghetti plot each line represents the change in the height (Figure 1) and BMI (Figure 2) for each participant in the study from birth to age 15 years. As expected, there was some variation in the rate of linear growth, but most participants clustered around the mean growth rates (represented by red line). Variability was more marked for BMI, with several participants showing marked deviation from the mean rates, mainly with higher BMI among females. Mean rates for linear growth and change in BMI in each growth period are shown in Figures S1 and S2 in the supplementary file. Annualized linear growth was fastest in the 0 – 6 months (early infancy) period, at 33.9 cm/year, and fell with each successive growth period to 5.2 cm/year in the 8 – 15 years growth period. BMI increased at a rate of 9 kg/m^2^ per year in the 0-6months period, but subsequently showed decrease in the 6-months to 2-years growth period and modest increase in the other growth periods. Spaghetti plots for systolic and diastolic BP measures at age 15 years and older are shown in Figures S3. There was marked variation in blood pressure intercept and slope with mean systolic blood pressure showing a slight decrease with age, while mean diastolic BP shown a modest increase with age.

**Table 1:** Participant characteristics at birth, during infancy, childhood, and adolescence.

| **Characteristic** | **Males**<br>**n = 162**<br>**Mean (SD)** | **Females**<br>**n = 203**<br>**Mean (SD)** | **Both Sexes**<br>**n = 365**<br>**Mean (SD)** |
| --- | --- | --- | --- |
| **Birth Characteristics** |  |  |  |
| Birth weight, [kg] | 3.25 (0.48) | 3.07 (0.51) | 3.15 (0.50) |
| Birth length, [cm] | 50.1 (2.68) | 49.1 (2.9) | 49.6 (2.8) |
| Gestation age, [weeks] | 39.3 (1.6) | 39.1 (1.9) | 39.2 (1.8) |
| BMI at birth [kg/m<sup>2</sup>] | 12.9 (1.5) | 12.7 (1.5) | 12.8 (1.5) |
| Preterm [% (n)] | 9.0 (14) | 11.2 (22) | 10.3 (36) |
| Low birth weight [% (n)] | 5.0 (8) | 11.9 (24) | 8.9 (32) |
| Socioeconomic status score | 19.7 (4.6) | 18.7 (4.3) | 19.1 (4.5) |
| **Characteristics at age 6 months** |  |  |  |
| Height, [cm] | 67.5 (2.3) | 65.7 (2.5) | 66.5 (2.6) |
| Weight, [kg] | 8.1 (1.0) | 7.4 (1.0) | 7.7 (1.0) |
| BMI, [kg/m<sup>2</sup>] | 17.6 (1.6) | 17.0 (1.7) | 17.3 (1.7) |
| Mean increase in height [cm] | 17.4 (2.3) | 16.6 (2.4) | 16.9 (2.4) |
| Mean increase in BMI [kg/m<sup>2</sup>] | 4.7 (1.8) | 4.3 (2.1) | 4.5 (2.0) |
| **Characteristics at age 2 years** |  |  |  |
| Height, [cm] | 87.8 (3.1) | 86.5 (3.3) | 87.1 (3.3) |
| Weight, [kg] | 12.5 (1.5) | 12.0 (1.6) | 12.2 (1.5) |
| BMI, [kg/m<sup>2</sup>] | 16.1 (1.3) | 15.9 (1.4) | 16.0 (1.3) |
| Mean increase in height, [cm] | 20.1 (2.0) | 20.8 (2.3) | 20.5 (2.2) |
| Mean increase in BMI [kg/m<sup>2</sup>] | -1.42 (1.2) | -1.10 (1.34) | -1.24 (1.3) |
| **Characteristics at age 8 years** |  |  |  |
| Height, [cm] | 131.1 (5.1) | 130.4 (5.5) | 130.7 (5.3) |
| Weight, [kg] | 29.7 (7.0) | 28.9 (6.8) | 29.2 (6.9) |
| BMI, [kg/m<sup>2</sup>] | 17.1 (3.1) | 16.9 (3.2) | 17.0 (3.2) |
| Mean increase in height, [cm] | 43.6 (3.2) | 43.9 (3.4) | 43.8 (3.3) |
| Mean increase in BMI, [kg/m<sup>2</sup>] | 1.08 (2.6) | 0.94 (2.7) | 1.00 (2.7) |
| **Characteristics at age 15 years** |  |  |  |
| Height, [cm] | 171.6 (6.7) | 163.2 (6.7) | 167.0 (7.9) |
| Weight, [kg] | 62.3 (15.2) | 59.3 (13.3) | 60.6 (14.2) |
| BMI, [kg/m<sup>2</sup>] | 21.0 (4.4) | 22.2 (4.7) | 21.7 (4.6) |
| Systolic BP, [mm Hg] | 114.4 (12.0) | 109.8 (10.5) | 111.9 (11.4) |
| Diastolic BP, [mm Hg] | 54.2 (7.4) | 53.5 (7.5) | 53.9 (7.4) |
| Mean increase in height, [cm] | 40.7 (3.9) | 32.9 (4.0) | 36.4 (5.5) |
| Mean increase in BMI, [kg/m<sup>2</sup>] | 4.06 (3.3) | 5.15 (3.2) | 4.65 (3.3) |

Reported values are means, with standard deviation (SD) in brackets except for preterm birth and low birth where the values are % (n).

**Figure 1:**
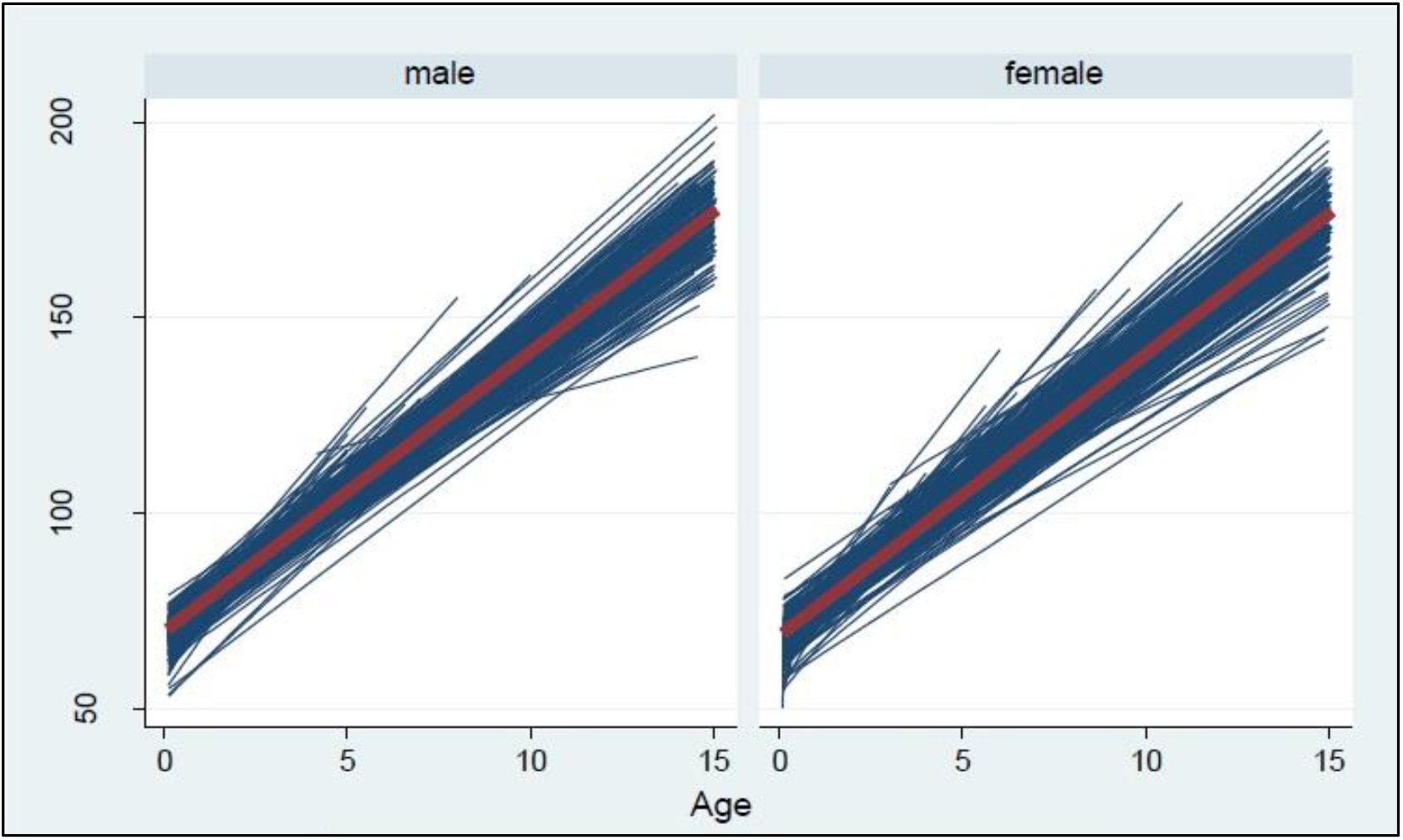
Spaghetti plot showing participants’ height (cm) vs. age (years) for male and female participants. Each line represents the change in the height (cm) for each participant in the study from birth to age 15 years. The centre red line represents the mean height for all participants together over the period.

**Figure 2:**
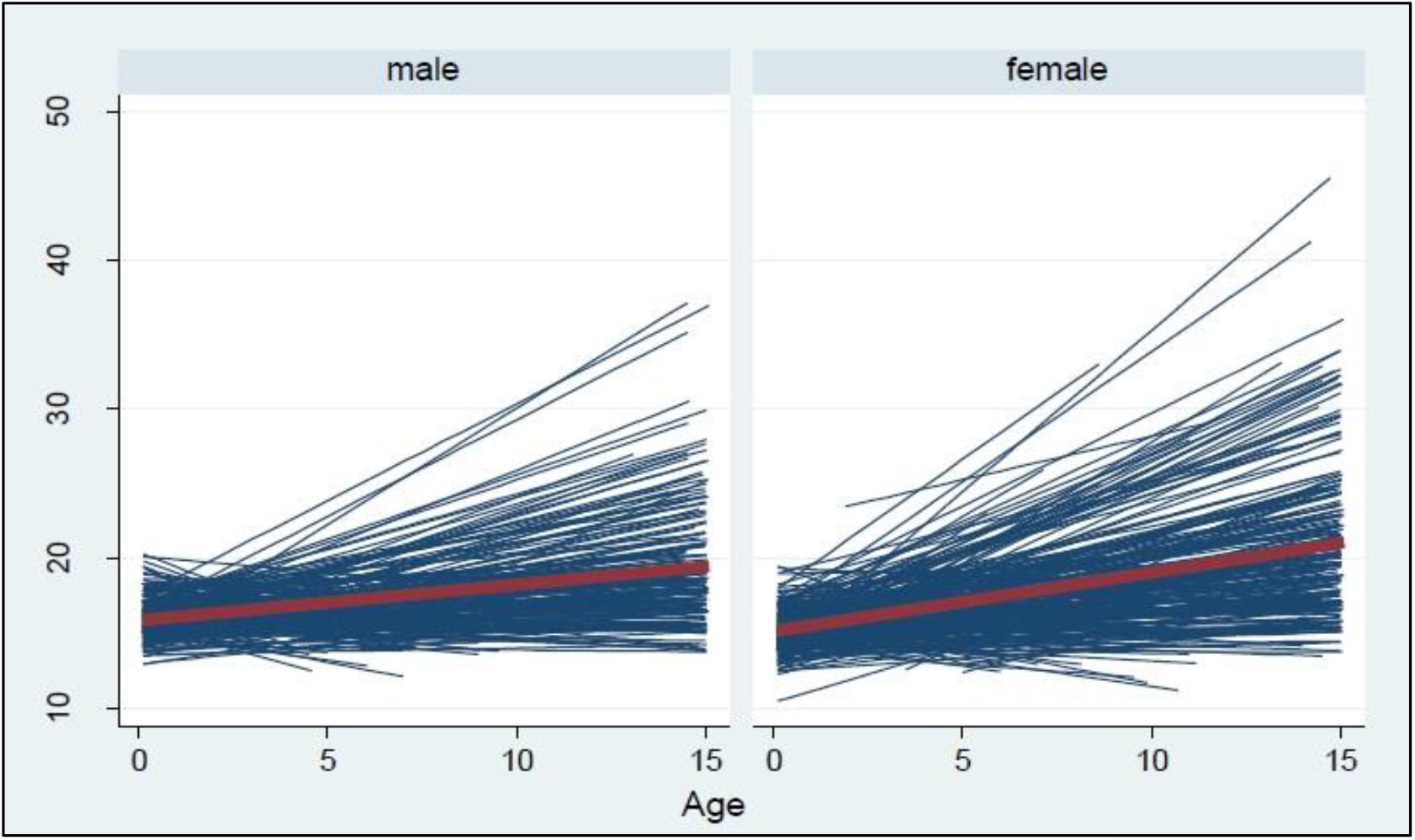
Spaghetti plot showing participants’ body mass index (BMI, kg/m^2^) vs. age (years) for male and female participants. Each line represents the change in the BMI (kg/m^2^) for each participant in the study from birth to age 15 years. The centre red line represents the mean BMI for all participants together over the period.

Regression coefficients for association between linear growth and SBP in sequentially adjusted models are shown in Table 2. Bivariate models are shown in Tables S3 and S4 in the supplementary file. In models adjusted for age and sex, there were significant positive associations between SBP and faster linear growth in the 0 - 6 months, 6 months - 2 years, and 2 - 8 years growth periods. When all the growth periods were included in the model these associations were mildly attenuated but remained statistically significant for the 0 - 6 months and 6 months - 2 years growth periods. Further adjustment for gestation age at birth and BMI at age 15 years further attenuated the coefficients, but the effect of linear growth in the 0-6 months period retained significance, while the effect for the 6 months – 2 years and 2-8 years now becoming only borderline significant. Male sex was also a significant predictor of higher SBP in this model, while older age was associated with lower SBP. Longer gestational age showed borderline significance for lower SBP.

**Table 2:** Regression coefficients for the effect of linear growth on systolic blood pressure at age 15 - 21 years.

| Variable | Regression Coefficient for SBP (mm Hg) | 95% Confidence Interval | P-value |
| --- | --- | --- | --- |
| **Linear growth within growth periods, adjusted for age & sex<sup>1</sup>** |  |  |  |
| Birth length [*z-score*] | 0.12 | -0.81, 1.06 | 0.793 |
| Linear growth 0-6 month | 1.46 | 0.70, 2.22 | <0.001 |
| Linear growth 6 months – 2 years | 1.10 | 0.24, 1.96 | 0.013 |
| Linear growth 2 years – 8 years | 0.88 | 0.10, 1.66 | 0.028 |
| Linear growth 8 years – 15 years | -0.43 | -1.52, 0.66 | <0.438 |
| **Model with all growth periods, adjusted for age and sex<sup>1</sup>** |  |  |  |
| Birth length (z-score) | 0.32 | -0.61, 1.25 | 0.502 |
| Linear growth 0-6 month | 1.21 | 0.31, 2.11 | 0.009 |
| Linear growth 6 months – 2 years | 0.92 | 0.12, 1.73 | 0.025 |
| Linear growth 2 years – 8 years | 0.85 | -0.04, 1.74 | 0.060 |
| Linear growth 8 years – 15 years | -0.41 | -1.50, 0.68 | 0.456 |
| Age [*years*] | -1.08 | -1.38, -0.78 | <0.001 |
| Sex [*male vs. female*] | 7.69 | 5.42, 9.96 | <0.001 |
| **Model with all growth periods adjusted for age, sex gestational age and BMI<sup>1</sup>** |  |  |  |
| Birth length [*z-score*] | 0.70 | -0.29, 1.69 | 0.166 |
| Linear growth rate 0-6 month | 1.06 | 0.26, 1.86 | 0.010 |
| Linear growth rate 6 months – 2 years | 0.74 | -0.07, 1.55 | 0.072 |
| Linear growth rate 2 years – 8 years | 0.66 | -0.06, 1.38 | 0.071 |
| Linear growth rate 8 years – 15 years | -0.09 | -0.94, 1.13 | 0.863 |
| Gestational age at birth [*days*] | -0.07 | -0.14, 0.01 | 0.071 |
| Age [*years*] | -1.09 | -1.38, -0.79 | <0.001 |
| Sex [*male vs. female*] | 7.31 | 5.19, 9.43 | <0.001 |
| BMI at age 15 years [*kg/m<sup>2</sup>*] | 0.37 | 0.18, 0.55 | <0.001 |

<sup>1</sup>Birth length and linear growth rate for each period are expressed as z-scores; coefficients represent the change in systolic blood pressure for a 1 standard deviation unit higher birth length or rate of change in in height for the growth period. The linear growth rates represent difference between the predicted height and observed height at end of the period based on the height at the start of the period.
Estimates obtained from 1909 systolic blood pressure measurements from 365 participants average of 5.2 readings per participant
BMI = Body mass index.

Similar models for DBP are shown in Table 3. Models adjusted for age and sex showed significant positive associations between DBP and faster linear growth in the 2 - 8 years growth period and borderline significance for the 0-6 months and 6 months - 2 years growth periods. In the model with all growth periods adjusted for age and sex, birth length became statistically significant, while there was borderline significance for the 0-6 months, 2-8 years, and 8-15 years growth periods. After further adjustment for gestational age at birth, current age, sex, and BMI at age 15 years none of the postnatal growth rates retained statistical significance, but birth length remained positively associated with higher DBP. Older age, and male sex were also positively associated with higher diastolic DBP.

**Table 3:**
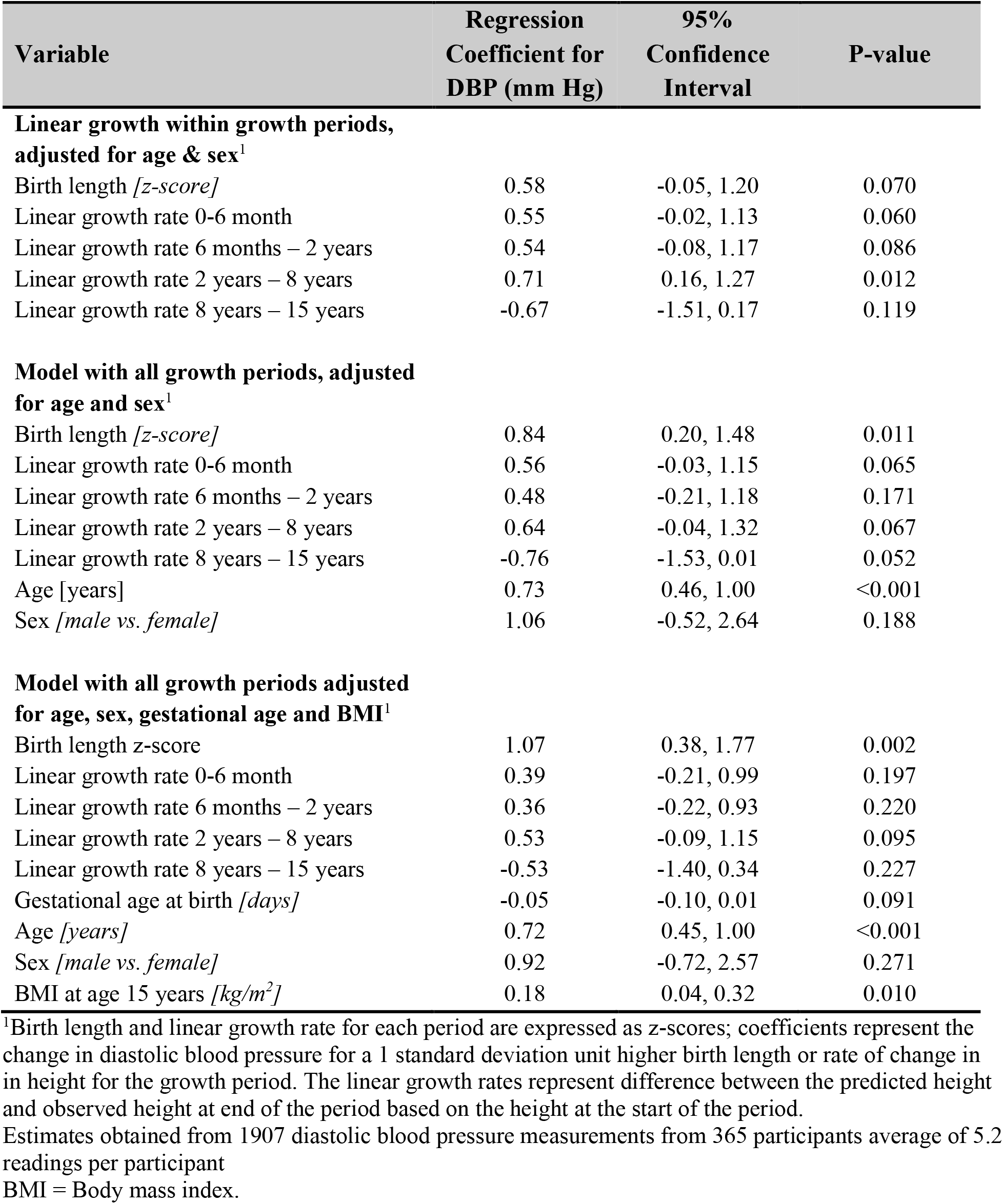
Regression coefficients for the effect of linear growth on diastolic blood pressure at age 15 - 21 years.

Models for the effect of rate of change in BMI on SBP are shown in Table 4. In the models adjusted for age and sex, significant positive associations were seen for higher SBP and greater rate of change in BMI in the 6-months to 2-years and 2 - 8 years and 8 - 15 years growth periods. There was little change in coefficients when change in BMI for all growth periods was included in the same model except that the 0 - 6 months growth period also became statistically significant. In the model with further adjustment for gestational age, birth length, current height and SES at birth, significant associations remained for the 6-months to 2-years, 2 – 8 years and 8-15 years growth periods.

**Table 4:**
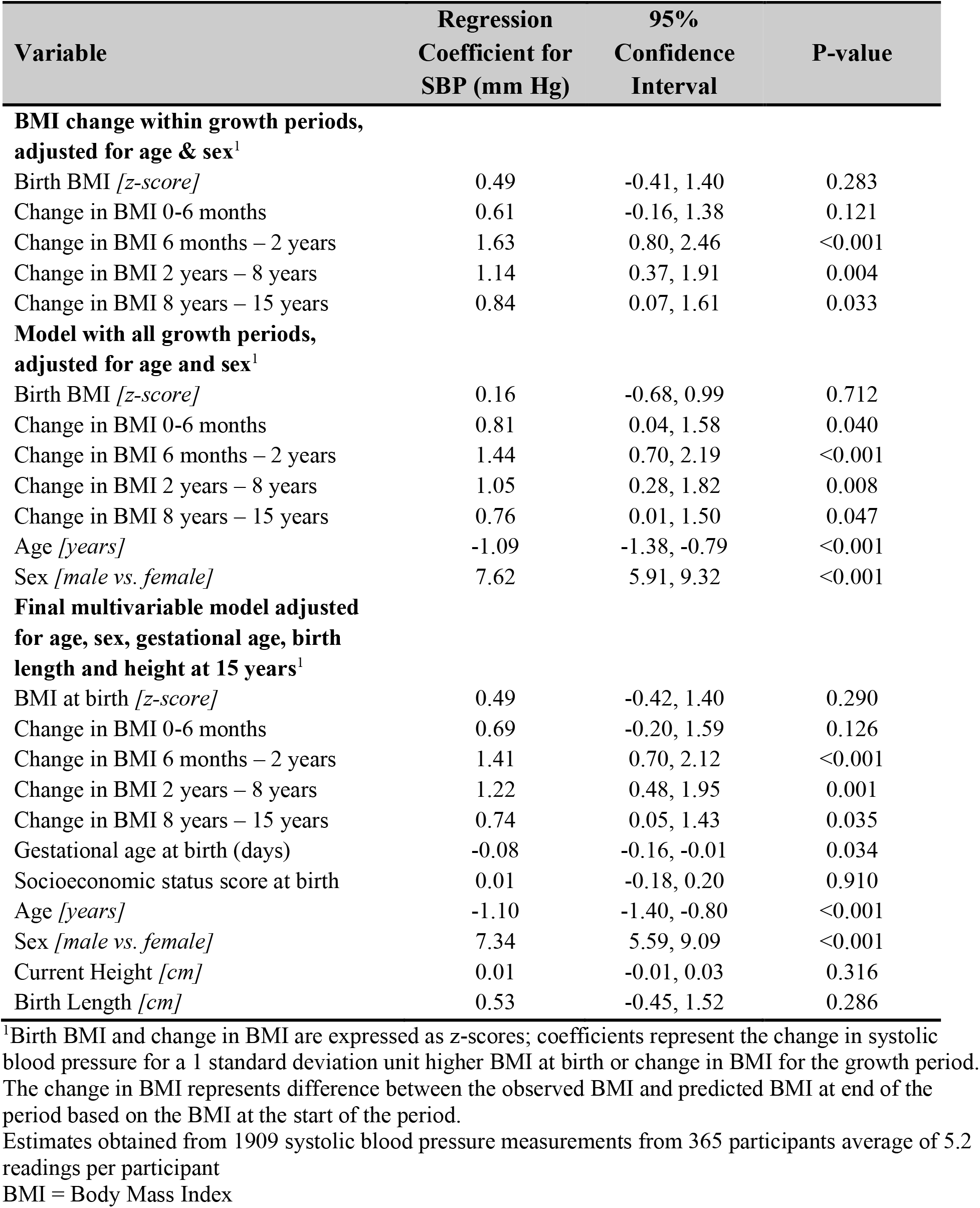
Regression coefficients for the effect of change in body mass index on systolic blood pressure at age 15 - 21 years.

Models for change in BMI and DBP are shown in Table 5. Significant positive associations were seen in models adjusted for age and sex for rate on change in BMI in the 6-months to 2- years, the 2 - 8 years, and 8 - 15 years growth periods. Again, only small changes in the coefficients were seen when all growth periods were modeled together. In the final models with further adjustment for age, sex, gestational age, current height and SES at birth, significant positive associations were seen for higher DBP with greater rate of increase in BMI for the 6-months to 2-years, 2 - 8 years and 8 - 15 years growth periods.

**Table 5:** Regression coefficients for the effect of change in body mass index on diastolic blood pressure at age 15 - 21 years.

| Variable | Regression Coefficient for DBP (mm Hg) | 95% Confidence Interval | P-value |
| --- | --- | --- | --- |
| **Linear growth within growth periods, adjusted for age & sex<sup>1</sup>** |  |  |  |
| Birth BMI [*z-score*] | 0.12 | -0.49, 0.73 | 0.705 |
| Change in BMI 0-6 months | 0.24 | -0.34, 0.83 | 0.416 |
| Change in BMI 6 months – 2 years | 0.78 | 0.17, 1.39 | 0.013 |
| Change in BMI 2 years – 8 years | 0.71 | 0.17, 1.25 | 0.011 |
| Change in BMI 8 years – 15 years | 0.62 | 0.01, 1.23 | 0.047 |
| **Model with all growth periods, adjusted for age and sex<sup>1</sup>** |  |  |  |
| Birth BMI [*z-score*] | -0.03 | -0.63, 0.58 | 0.934 |
| Change in BMI 0-6 month | 0.32 | -0.28, 0.91 | 0.291 |
| Change in BMI 6 months – 2 years | 0.64 | 0.06, 1.23 | 0.031 |
| Change in BMI 2 years – 8 years | 0.64 | 0.05, 1.23 | 0.033 |
| Change in BMI 8 years – 15 years | 0.72 | 0.11, 1.32 | 0.021 |
| Age [*years*] | 0.73 | 0.45, 1.00 | <0.001 |
| Sex [*male vs. female*] | 0.53 | -0.68, 1.75 | 0.392 |
| **Model with all growth periods adjusted for age, sex, gestational age, and birth length height<sup>1</sup>** |  |  |  |
| Birth BMI [*z-score*] | 0.13 | -0.50, 0.76 | 0.690 |
| Change in BMI 0-6 month | 0.33 | -0.23, 0.88 | 0.251 |
| Change in BMI 6 months – 2 years | 0.63 | 0.10, 1.17 | 0.021 |
| Change in BMI 2 years – 8 years | 0.62 | 0.07, 1.18 | 0.028 |
| Change in BMI 8 years – 15 years | 0.66 | 0.12, 1.20 | 0.016 |
| Gestational age at birth [*days*] | -0.04 | -0.10, 0.01 | 0.121 |
| Socioeconomic status score at birth [*sum score*] | 0.05 | -0.08, 0.19 | 0.432 |
| Age [*years*] | 0.72 | 0.45, 0.99 | <0.001 |
| Sex [*male vs. female*] | 0.21 | -1.03, 1.45 | 0.740 |
| Current Height [*cm*] | -0.005 | -0.02, 0.13 | 0.602 |
| Birth length [*z-score*] | 0.83 | 0.16, 1.50 | 0.015 |

<sup>1</sup>Birth BMI and change in BMI are expressed as z-scores; coefficients represent the change in systolic blood pressure for a 1 standard deviation unit higher BMI at birth or change in BMI for the growth period. The change in BMI represents the predicted BMI and observed at end of the period based on the BMI at the start of the period.
Estimates obtained from 1907 diastolic blood pressure measurements from 365 participants average of 5.2 readings per participant

We also assessed the combined effects of linear growth and change in adiposity on SBP and DBP by including the conditional growth variables for both BMI and linear growth in the same models. The results for these models are shown in Table 6. There was little correlation between these variables, with correlation coefficients generally <0.4 as shown in Table S1 in the supplementary file. For SBP only linear growth in the 0-6 months period was associated with higher SBP, while greater change in BMI in the 6 months to 2-years, 2 - 8 years and the 8-15 years remained statistically significant. For diastolic BP only change in BMI in the 8-15 years period remained significant.

**Table 6:** Regression coefficients for the combined effect of linear growth and change in body mass index on systolic and diastolic blood pressure at age 15-21 years.

| Variable | Regression<br>Coefficient for<br>DBP (mm Hg) | 95%<br>Confidence<br>Interval | P-value |
| --- | --- | --- | --- |
| **Systolic Blood Pressure<sup>1</sup>** |  |  |  |
| Linear growth rate 0-6 months | 1.00 | 0.22, 1.77 | 0.011 |
| Linear growth rate 6 months – 2 years | 0.70 | -0.25, 1.66 | 0.145 |
| Linear growth rate 2 years – 8 years | 0.61 | -0.20, 1.42 | 0.136 |
| Linear growth rate 8 years – 15 years | 0.36 | -0.80, 1.51 | 0.593 |
| Change in BMI 0-6 months | 0.44 | -0.40, 1.28 | 0.305 |
| Change in BMI 6 months – 2 years | 1.47 | 0.62, 2.32 | 0.001 |
| Change in BMI 2 years – 8 years | 0.88 | 0.06, 1.70 | 0.035 |
| Change in BMI 8 years – 15 years | 0.81 | 0.04, 1.58 | 0.040 |
| Age | -1.08 | -1.37, -0.78 | <0.001 |
| Sex (male vs. female) | 7.01 | 4.59, 9.42 | <0.01 |
| **Diastolic Blood Pressure<sup>1</sup>** |  |  |  |
| Linear growth rate 0-6 month | 0.48 | -0.18, 1.13 | 0.151 |
| Linear growth rate 6 months – 2 years | 0.27 | -0.46, 0.99 | 0.469 |
| Linear growth rate 2 years – 8 years | 0.44 | -0.20, 1.08 | 0.174 |
| Linear growth rate 8 years – 15 years | -0.34 | -1.31, 0.63 | 0.483 |
| Change in BMI 0-6 months | 0.13 | -0.44, 0.70 | 0.660 |
| Change in BMI 6 months – 2 years | 0.54 | -0.10, 1.19 | 0.100 |
| Change in BMI 2 years – 8 years | 0.32 | -0.33, 0.98 | 0.328 |
| Change in BMI 8 years – 15 years | 0.63 | 0.02, 1.25 | 0.044 |
| Age (years) | 0.73 | 0.46, 1.00 | <0.001 |
| Sex (male vs. female) | 0.92 | -0.90, 2.74 | 0.320 |

<sup>1</sup>Both linear growth and change in BMI are expressed as z-scores; coefficients represent the change in systolic or diastolic blood pressure for a 1 standard deviation unit higher value for the growth period. The linear growth and change in BMI represent the difference between the predicted value and observed value at end of the period based on the value at the start of the period.
Estimates based on 1909 systolic blood pressure measurements and 1907 diastolic blood pressure measurements from 365 participants with an average of 5.2 readings per participant

In order to determine if the observed effects differed among persons born with low birth weight, we ran additional models stratified by birth weight category. Details for these models are shown in Tables S5 and S6 in the supplementary file. Among persons with low birth weight, birth BMI showed a statistically significant inverse association with both systolic and diastolic BP, whereas among persons with normal birthweight there was a non-significant positive association. For the linear growth rates and change in BMI variables, the strength and direction of association were generally similar for both persons with low birth weight and normal birth weight; however, several of the estimates among persons with low birth weight were not statistically significant, given the much smaller number of observations.

## DISCUSSION

We have shown that both faster linear growth rate and greater rate of increase in BMI during childhood are associated with higher systolic and diastolic BP. The effect of faster linear growth was strongest in early infancy (0-6 months) for systolic blood pressure, and in the 2-8 years growth period for diastolic blood pressure. For change in BMI, the effects were strongest in the late infancy (6 months – 2 years) and early childhood (2-8 years) growth period for SBP but was similar for all growth periods from after early infancy for DBP. When both linear growth and change in BMI were included in the same model faster linear growth was significantly associated with SBP only in the 0-6 months growth period, while greater gain in BMI for the three growth periods from 6 months to 15 years was associated with higher SBP and only greater increase in BMI in the 8-15 years growth period was associated with higher DBP.

Our findings add support to the literature showing an effect of postnatal growth on BP; in particular, the effects of linear growth in early infancy and increasing obesity throughout childhood. That these data are from an Afro-Caribbean population in a developing country setting increases its relevance, as many of the previous studies were done among Caucasian populations in developed countries. The findings in this study are generally consistent with the published literature showing that faster increase in both height and weight are associated with higher SBP and DBP in later childhood or early adulthood (17, 18, 36-40). The majority of studies focused on infancy or early childhood, but some also included later childhood and adolescent growth (36, 39). The effect size in our study was also similar to the findings in the other studies with a 1 standard deviation unit increase in growth rate resulting in a 1-2 mm Hg increase in SBP and 0.5-1 mm Hg increase in DBP. Consistent with the findings in our study, other studies have also found that the effect of childhood or adolescent growth was attenuated and sometimes no longer significant after adjustment for body size at the time of BP assessment (36, 40). Taking the findings of this and the other studies cited together, there is therefore strong evidence that faster than expected growth in childhood and adolescence is associated with higher BP in later childhood and early adulthood and thus confers a more adverse cardiovascular risk profile. Our study suggests that except for linear growth in the 0-6 months period gain in BMI has a stronger effect on BP in late adolescence or early adulthood.

It should be noted that we did not find an inverse relationship between birth weight and SBP or DBP in this study. The lack of association with birth weight has been reported in other studies (16, 41), but the overwhelming body of evidence, including other local studies, supports the inverse association between birth weight and blood pressure (26-28, 42-44). The absence of an association here may be related to the small sample size, and thus a chance occurrence.

Possible mechanisms underlying the association between faster postnatal growth and higher BP include its association with greater levels of obesity in adulthood as well as physiological programming of the cardiovascular system. The role of adult BMI is supported by the attenuation of the growth effects when adjusting for current body size as seen in this and other studies.

Studies suggest that cardiovascular development remains susceptible to environmental perturbations in early childhood, thus faster than expected growth may program arterial physiology through alteration in the deposition of elastin and collagen in the arterial wall, or through alterations in the hypothalamo-pituitary adrenal axis, and the sympathetic nervous system (17, 37, 45-47). Singhal and Lucus (46) have proposed the ‘growth acceleration hypothesis’, which suggests that adverse long-term effect of faster growth may be the common factor in the programming of cardiovascular disease. Epigenetic modification is now thought to play a central role in the developmental origins of disease (1, 8).

With regards to public health importance there is still much debate as to the implications of the association between faster postnatal growth and adverse cardiovascular health (19, 23). Adair and colleagues have suggested that while faster postnatal growth may be associated with poorer cardiovascular health, there are potential gains, in particular, in reducing adverse childhood outcomes and improvements in social capital, specifically reducing short stature and improved schooling (10). They argue that it may be best to promote optimal linear growth in the first two year of life and avoidance of excessive weight gain afterwards. Our data would support this position. These recommendations are also consistent with those of Jain & Singhal (23), who recommend close monitoring of weight gain and discouraging overfeeding and crossing of centiles. We believe that these recommendations would be applicable in our setting as well.

Strengths of this study include the fact that we have used prospectively collected data with several measurements of body size and blood pressure, using standardized protocols. The use of conditional measurements of linear growth and BMI mitigated the impact of the problems of collinearity between serial measurements of body size thus allowing us to include several growth periods in the simultaneously in the assessment. Additionally, we included growth measurements from the birth to adolescence, unlike several studies which only included early childhood measurements.

Limitations of this study include its relatively small size and the fact that there was some degree of attrition from the original cohort over time. Participants included in the analysis were however similar to the full cohort except for a marginally higher birth weight. The availability of several BP measures over an average of 5 visits after age 15 years provided a much larger number of BP measurements which partially compensated for the small sample size. Some participants also had missing data for some measurements. We were able to overcome the challenge of missing data by using the multiple imputation to fill in missing values. Another potential limitation is the use of BMI as the main measure of body fatness. We note that while BMI is a good measure of body fatness, it does not distinguish between fat mass and muscle mass (48). However, BMI has been used in many epidemiological studies and has high correlation with cardiovascular outcomes, thus despite its limitations, it is useful in clinical and epidemiological studies (49).

This is the first study to evaluate the role of postnatal growth and blood pressure beyond childhood in an Afro-Caribbean population. Walker and colleagues (27) looked at the effect of postnatal growth and blood pressure in a cohort of stunted and non-stunted children, but children were only 11-12 years old at the time of assessment. Previous analyses from this cohort were also limited to periods from birth up to 14 years old (25, 29). This study was population based, with generally healthy participants, suggesting that the findings can be generalized to the Jamaican population. We acknowledge that loss to follow up (approximately 36%) may limit generalizability of this study, but our findings still represent the best available evidence for our population. Additionally, the associations being studied were more biological characteristics, so there was little reason to suspect that the effect seen in the studied sample would be largely different from the Jamaican population. This paper therefore is an important addition to the literature and will contribute to policy initiatives with regards to nutrition practice recommendation in Afro-Caribbean and similar populations.

## CONCLUSION

This study found that both faster linear growth and greater rate of increase in BMI were associated with higher SBP and DBP in Afro-Caribbean youth. The association was most consistent for linear growth in the early infancy and for faster gain in BMI from late infancy to adolescence. These findings suggest that postnatal growth trajectories should be closely monitored and faster than expected gain in BMI should be discouraged.

## Supporting information

STROBE Checklist

## Data Availability

Data used in this manuscript are available from the authors upon request.

## ACKNOWLEDGMENTS

The authors acknowledge with thanks the project staff (nurses, laboratory personnel, administrative staff, and project assistants) for their contribution to the project. We also thank the funding agencies which sponsored this study.

## AUTHOR CONTRIBUTIONS

**TSF** – conceptualized paper, developed data-analysis plan along with TRT, interpreted data, wrote first draft of manuscript, and revised the manuscript after critical reviewed by coauthors.

**TRT** - lead the data analysis, contributed to interpretation of data, and critically reviewed the manuscript

**LCH** – contributed to data collection and critical review of the manuscript

**MMT** – contributed to the design of original study, data collection, interpretation of data and critically review of manuscript

**TEF-** lead design of original study, contributed to data collection and critical review of the manuscript

**CO** – contributed to the design of original study, interpretation of data, statistical review, and critical review of the manuscript

**MSB -** contributed to the conceptualization of the paper, interpretation of data and critical review of the manuscript

**RJW** – contributed to the design of original study, conceptualization of the paper, interpretation of data, and critical review of the manuscript

## REFERENCES

1. Baird J, Jacob C, Barker M, Fall CH, Hanson M, Harvey NC, et al. Developmental Origins of Health and Disease: A Lifecourse Approach to the Prevention of Non-Communicable Diseases. Healthcare (Basel). 2017;5(1).

2. Barker DJ. The developmental origins of adult disease. J AmCollNutr. 2004;23(6 Suppl):588S–95S.

3. Gluckman PD, Buklijas T, Hanson MA. The Developmental Origins of Health and Disease (DOHaD) Concept: Past, Present, and Future. The Epigenome and Developmental Origins of Health and Disease. Boston: Academic Press; 2016. p. 1–15.

4. Murray R, Godfrey KM, Lillycrop KA. The Early Life Origins of Cardiovascular Disease. Curr Cardiovasc Risk Rep. 2015;9(4):15.

5. Barker DJP, Osmond C, Forsen TJ, Kajantie E, Eriksson JG. Maternal and Social Origins of Hypertension. Hypertension. 2007;50(3):565–71.

6. Lawlor DA, Smith GD. Early life determinants of adult blood pressure. Curr Opin Nephrol Hypertens. 2005;14(3):259–64.

7. Whincup PH, Cook DG, Geleijnse JH. A life course approach to blood pressure. In: Kuh D, Ben-Shlomo Y, editors. A life course approach to chronic disease epidemiology. New York: Oxford University Press; 2004. p. 218–39.

8. Hanson MA, Gluckman PD. Early developmental conditioning of later health and disease: physiology or pathophysiology? Physiol Rev. 2014;94(4):1027–76.

9. Morton JS, Cooke CL, Davidge ST. In Utero Origins of Hypertension: Mechanisms and Targets for Therapy. Physiol Rev. 2016;96(2):549–603.

10. Adair LS, Fall CH, Osmond C, Stein AD, Martorell R, Ramirez-Zea M, et al. Associations of linear growth and relative weight gain during early life with adult health and human capital in countries of low and middle income: findings from five birth cohort studies. Lancet. 2013;382(9891):525–34.

11. Barker DJ, Osmond C, Forsen TJ, Kajantie E, Eriksson JG. Trajectories of growth among children who have coronary events as adults. N Engl J Med. 2005;353(17):1802–9.

12. Bhargava SK, Sachdev HS, Fall CH, Osmond C, Lakshmy R, Barker DJ, et al. Relation of serial changes in childhood body-mass index to impaired glucose tolerance in young adulthood. N Engl J Med. 2004;350(9):865–75.

13. Bowers K, Liu G, Wang P, Ye T, Tian Z, Liu E, et al. Birth weight, postnatal weight change, and risk for high blood pressure among chinese children. Pediatrics. 2011;127(5):e1272–9.

14. Eriksson JG, Forsen TJ, Kajantie E, Osmond C, Barker DJ. Childhood growth and hypertension in later life. Hypertension. 2007;49(6):1415–21.

15. Halldorsson TI, Gunnarsdottir I, Birgisdottir BE, Gudnason V, Aspelund T, Thorsdottir I. Childhood growth and adult hypertension in a population of high birth weight. Hypertension. 2011;58(1):8–15.

16. Leunissen RW, Kerkhof GF, Stijnen T, Hokken-Koelega AC. Effect of birth size and catch-up growth on adult blood pressure and carotid intima-media thickness. Horm Res Paediatr. 2012;77(6):394–401.

17. Perng W, Rifas-Shiman SL, Kramer MS, Haugaard LK, Oken E, Gillman MW, et al. Early Weight Gain, Linear Growth, and Mid-Childhood Blood Pressure: A Prospective Study in Project Viva. Hypertension. 2016;67(2):301–8.

18. Howe LD, Chaturvedi N, Lawlor DA, Ferreira DLS, Fraser A, Davey Smith G, et al. Rapid increases in infant adiposity and overweight/obesity in childhood are associated with higher central and brachial blood pressure in early adulthood. J Hypertens. 2014;32(9):1789–96.

19. Hales CN, Ozanne SE. The dangerous road of catch-up growth. J Physiol. 2003;547(Pt 1):5–10.

20. Ong KK. Catch-up growth in small for gestational age babies: good or bad? Curr Opin Endocrinol Diabetes Obes. 2007;14(1):30–4.

21. Ong KK, Ahmed ML, Emmett PM, Preece MA, Dunger DB. Association between postnatal catch-up growth and obesity in childhood: prospective cohort study. BMJ. 2000;320(7240):967–71.

22. Victora CG, Barros FC, Horta BL, Martorell R. Short-term benefits of catch-up growth for small-for-gestational-age infants. Int J Epidemiol. 2001;30(6):1325–30.

23. Jain V, Singhal A. Catch up growth in low birth weight infants: striking a healthy balance. Rev Endocr Metab Disord. 2012;13(2):141–7.

24. Boyne MS, Thompson DS, Osmond C, Fraser RA, Thame MM, Taylor-Bryan C, et al. The effect of antenatal factors and postnatal growth on serum adiponectin levels in children. J Dev Orig Health Dis. 2013;4(4):317–23.

25. Boyne MS, Osmond C, Fraser RA, Reid M, Taylor-Bryan C, Soares-Wynter S, et al. Developmental origins of cardiovascular risk in Jamaican children: the Vulnerable Windows Cohort study. Br J Nutr. 2010;104(7):1026–33.

26. Forrester TE, Wilks RJ, Bennett FI, Simeon D, Osmond C, Allen M, et al. Fetal growth and cardiovascular risk factors in Jamaican schoolchildren. BMJ. 1996;312(7024):156.

27. Walker SP, Gaskin P, Powell CA, Bennett FI, Forrester TE, Grantham-McGregor S. The effects of birth weight and postnatal linear growth retardation on blood pressure at age 11-12 years. J Epidemiol Community Health. 2001;55(6):394–8.

28. Ferguson TS, Younger-Coleman NO, Tulloch-Reid MK, Knight-Madden JM, Bennett NR, Samms-Vaughan M, et al. Birth weight and maternal socioeconomic circumstances were inversely related to systolic blood pressure among Afro-Caribbean young adults. J Clin Epidemiol. 2015;68(9):1002–9.

29. Royal-Thomas T, McGee D, Sinha D, Osmond C, Forrester T. Association of maternal blood pressure in pregnancy with blood pressure of their offspring through adolescence. J Perinat Med. 2015;43(6):695–701.

30. Boyne MS, Thame M, Osmond C, Fraser RA, Gabay L, Reid M, et al. Growth, body composition, and the onset of puberty: longitudinal observations in Afro-Caribbean children. JClinEndocrinolMetab. 2010;95(7):3194–200.

31. Thame M, Osmond C, Wilks RJ, Bennett FI, McFarlane-Anderson N, Forrester TE. Blood pressure is related to placental volume and birth weight. Hypertension. 2000;35(2):662–7.

32. Cole TJ, Henson GL, Tremble JM, Colley NV. Birthweight for length: ponderal index, body mass index or Benn index? Ann Hum Biol. 1997;24(4):289–98.

33. Lohr SL. Sampling: Design and Analysis. Pacific Grove, CA: Duxbury Press; 1999.

34. Kajantie E, Barker DJ, Osmond C, Forsen T, Eriksson JG. Growth before 2 years of age and serum lipids 60 years later: the Helsinki Birth Cohort study. Int J Epidemiol. 2008;37(2):280–9.

35. Marshall A, Altman DG, Holder RL, Royston P. Combining estimates of interest in prognostic modelling studies after multiple imputation: current practice and guidelines. BMC Med Res Methodol. 2009;9:57.

36. Adair LS, Martorell R, Stein AD, Hallal PC, Sachdev HS, Prabhakaran D, et al. Size at birth, weight gain in infancy and childhood, and adult blood pressure in 5 low-and middle-income-country cohorts: when does weight gain matter? Am J Clin Nutr. 2009;89(5):1383–92.

37. Ben-Shlomo Y, McCarthy A, Hughes R, Tilling K, Davies D, Smith GD. Immediate postnatal growth is associated with blood pressure in young adulthood: the Barry Caerphilly Growth Study. Hypertension. 2008;52(4):638–44.

38. Jones A, Charakida M, Falaschetti E, Hingorani AD, Finer N, Masi S, et al. Adipose and height growth through childhood and blood pressure status in a large prospective cohort study. Hypertension. 2012;59(5):919–25.

39. Sinaiko AR, Donahue RP, Jacobs DR, Jr., Prineas RJ. Relation of weight and rate of increase in weight during childhood and adolescence to body size, blood pressure, fasting insulin, and lipids in young adults. The Minneapolis Children’s Blood Pressure Study. Circulation. 1999;99(11):1471–6.

40. Law CM, Shiell AW, Newsome CA, Syddall HE, Shinebourne EA, Fayers PM, et al. Fetal, Infant, and Childhood Growth and Adult Blood Pressure: A Longitudinal Study From Birth to 22 Years of Age. Circulation. 2002;105(9):1088–92.

41. Menezes AM, Hallal PC, Horta BL, Araujo CL, Vieira Mde F, Neutzling M, et al. Size at birth and blood pressure in early adolescence: a prospective birth cohort study. Am J Epidemiol. 2007;165(6):611–6.

42. Gamborg M, Byberg L, Rasmussen F, Andersen PK, Baker JL, Bengtsson C, et al. Birth Weight and Systolic Blood Pressure in Adolescence and Adulthood: Meta-Regression Analysis of Sex- and Age-specific Results from 20 Nordic Studies. Am J Epidemiol. 2007;166(6):634–45.

43. Huxley RR, Shiell AW, Law CM. The role of size at birth and postnatal catch-up growth in determining systolic blood pressure: a systematic review of the literature. J Hypertens. 2000;18(7):815–31.

44. Mu M, Wang SF, Sheng J, Zhao Y, Li HZ, Hu CL, et al. Birth weight and subsequent blood pressure: a meta-analysis. Arch Cardiovasc Dis. 2012;105(2):99–113.

45. Martyn CN, Greenwald SE. Impaired synthesis of elastin in walls of aorta and large conduit arteries during early development as an initiating event in pathogenesis of systemic hypertension. Lancet. 1997;350(9082):953–5.

46. Singhal A, Lucas A. Early origins of cardiovascular disease: is there a unifying hypothesis? Lancet. 2004;363(9421):1642–5.

47. Boyne MS, Woollard A, Phillips DIW, Taylor-Bryan C, Bennett FI, Osmond C, et al. The association of hypothalamic-pituitary-adrenal axis activity and blood pressure in an Afro-Caribbean population. Psychoneuroendocrinology. 2009;34(5):736–42.

48. Weber DR, Moore RH, Leonard MB, Zemel BS. Fat and lean BMI reference curves in children and adolescents and their utility in identifying excess adiposity compared with BMI and percentage body fat. Am J Clin Nutr. 2013;98(1):49–56.

49. Center for Disease Control. Body Mass Index: Consideration for Practioners2011 September 16, 2020. Available from: https://www.cdc.gov/obesity/downloads/BMIforpactitioners.pdf.

