## Supplementary Tables for "EFFECT OF LINEAR GROWTH RATE AND CHANGE IN BODY MASS INDEX IN CHILDHOOD AND ADOLESCENCE ON BLOOD PRESSURE IN AFRO-CARIBBEAN YOUTH: THE VULNERABLE WINDOWS COHORT STUDY"

### Supplementary Tables and Figures

**Table S1: Correlation coefficients for linear growth and rate of change BMI together at given time periods**

| Variable | cH 0-6 months | cH 6 months – 2 years | cH 2-8 years | cH 8-15 years | cBMI 0-6 months | cBMI 6 months – 2 years | cBMI 2-8 years | cBMI 8-15 years |
|---|---|---|---|---|---|---|---|---|
| **cH 0-6 months** | 1.0 | | | | | | | |
| **cH 6 months – 2 years** | 0.076 | 1.0 | | | | | | |
| **cH 2-8 years** | 0.095 | 0.040 | 1.0 | | | | | |
| **cH 8-15 years** | 0.268 *** | -0.177 * | -0.227 *** | 1.0 | | | | |
| **cBMI 0-6 months** | 0.235 *** | 0.147 * | -0.003 | 0.119 | 1.0 | | | |
| **cBMI 6 months – 2 years** | 0.058 | 0.155 ** | 0.007 | -0.193 ** | -0.049 | 1.0 | | |
| **cBMI 2-8 years** | 0.025 | 0.199 ** | 0.330 *** | -0.229 *** | -0.105 | 0.144 * | 1.0 | |
| **cBMI 8-15 years** | -0.024 | -0.069 | 0.035 | -0.111 | -0.022 | 0.014 | -0.018 | 1.0 |

Coefficients are derived from Person's correlation. All variables were entered as z-score; cH = change in height; cBMI = change in body mass index.

*p<0.05; **p<0.01; ***p<0.001

**Table S2: Number of missing and non-missing values for variables used in the analysis**

| Variable | Number of missing observations | Number of non-missing observations | % Missing |
|---|---|---|---|
| Birth Weight | 28 | 1879 | 1.5 |
| Birth length | 45 | 1862 | 2.4 |
| Birth BMI | 45 | 1862 | 2.4 |
| Gestational age at birth | 55 | 1852 | 2.9 |
| Socioeconomic status | 1 | 1906 | 0.1 |
| Change in BMI 0-6 month | 291 | 1616 | 15.3 |
| Change in BMI 6 months – 2 years | 396 | 1511 | 20.8 |
| Change in BMI 2 years – 8 years | 492 | 1415 | 25.8 |
| Change in BMI 8 years – 15 years | 468 | 1439 | 24.5 |
| Linear growth rate 0-6 months | 291 | 1616 | 15.3 |
| Linear growth rate 6 months – 2 years | 396 | 1511 | 20.8 |
| Linear growth rate 2 years – 8 years | 492 | 1415 | 25.8 |
| Linear growth rate 8 years – 15 years | 465 | 1442 | 24.4 |

Analyses were limited to participants with available blood pressure measurements after age 15 years. Estimates based on 1907 estimates used for diastolic BP models. Systolic blood pressure used 1909 observations. There were no missing values for age or sex among participants included in the analysis.

**Table S3: Bivariate models for systolic blood pressure from age 15-21 years as outcome variable**

| Variable | Regression Coefficient | 95% Confidence Interval | P-value |
|---|---|---|---|
| Birth length z-score | 0.66 | -0.31, 1.64 | 0.182 |
| Linear growth rate 0-6 month | 2.06 | 1.28, 2.84 | <0.001 |
| Linear growth rate 6 months – 2 years | 0.72 | -0.23, 1.68 | 0.138 |
| Linear growth rate 2 years – 8 years | 0.65 | -0.11, 1.42 | 0.091 |
| Linear growth rate 8 years – 15 years | 1.54 | 0.74, 2.34 | <0.001 |
| Age (years) | -1.10 | -1.40, -0.80 | <0.001 |
| Sex (male vs. female) | 7.43 | 5.68, 9.18 | <0.001 |
| Height at age 15 | 0.08 | 0.02, 0.14 | 0.009 |
| BMI at age 15 | 0.32 | 0.13, 0.52 | 0.001 |
| Birth weight | 0.90 | -0.07, 1.87 | 0.069 |
| Birth length | 0.66 | -0.31, 1.64 | 0.182 |
| Gestational age (days) | -0.04 | -0.12, 0.03 | 0.247 |
| Socioeconomic status at birth | 0.09 | -0.12, 0.31 | 0.390 |

**Table S4: Bivariate models for diastolic blood pressure from age 15-21 years as outcome variable**

| Variable | Regression Coefficient | 95% Confidence Interval | P-value |
|---|---|---|---|
| Birth length z-score | 0.63 | 0.01, 1.26 | 0.047 |
| Linear growth rate 0-6 month | 0.54 | 0.01, 1.06 | 0.045 |
| Linear growth rate 6 months – 2 years | 0.49 | -0.08, 1.06 | 0.092 |
| Linear growth rate 2 – 8 years | 0.59 | 0.02, 1.17 | 0.044 |
| Linear growth rate 8 – 15 years | -0.19 | -0.77, 0.39 | 0.514 |
| Age (years) | 0.72 | 0.45, 0.99 | <0.001 |
| Sex (male vs. female) | 0.43 | -0.78, 1.65 | 0.485 |
| Height at age 15 | 0.01 | -0.03, 0.05 | 0.705 |
| BMI at age 15 | 0.22 | 0.09, 0.36 | 0.001 |
| Birth weight | 0.53 | -0.07, 1.12 | 0.082 |
| Birth length | 0.63 | 0.01, 1.26 | 0.047 |
| Gestational age (days) | -0.01 | -0.06, 0.04 | 0.646 |
| Socioeconomic status at birth | 0.04 | -0.09, 0.18 | 0.517 |

**Table S5: Stratified models for postnatal linear growth and systolic and diastolic blood pressure by birth weight category**

| Variable | Low Birth Weight | | Normal Birth Weight | |
|---|---|---|---|---|
| | Regression Coefficient | P-value | Regression Coefficient | P-value |
| **Linear Growth with SBP** | | | | |
| Birth length z-score | 1.00 | 0.578 | 0.56 | 0.323 |
| Linear growth rate 0-6 months | 1.52 | 0.264 | 1.09 | 0.015 |
| Linear growth rate 6 months – 2 years | 1.09 | 0.288 | 0.70 | 0.116 |
| Linear growth rate 2 years – 8 years | -0.04 | 0.975 | 0.58 | 0.167 |
| Linear growth rate 8 years – 15 years | 0.71 | 0.652 | -0.07 | 0.916 |
| Gestational age at birth (days) | 0.02 | 0.826 | -0.11 | 0.013 |
| Age (years) | -0.42 | 0.435 | -1.13 | <0.001 |
| Sex (male vs. female) | 7.66 | 0.020 | 7.24 | <0.001 |
| BMI at age 15 ($kg/m^2$) | 0.48 | 0.091 | 0.33 | 0.001 |
| **Linear Growth with DBP** | | | | |
| Birth length z-score | 1.24 | 0.355 | 1.04 | 0.009 |
| Linear growth rate 0-6 months | 0.76 | 0.560 | 0.36 | 0.261 |
| Linear growth rate 6 months – 2 years | 0.05 | 0.956 | 0.45 | 0.150 |
| Linear growth rate 2 years – 8 years | -0.36 | 0.759 | 0.57 | 0.082 |
| Linear growth rate 8 years – 15 years | -0.65 | 0.704 | -0.65 | 0.213 |
| Gestational age at birth (days) | -0.02 | 0.694 | -0.06 | 0.060 |
| Age (years) | 1.59 | 0.001 | 0.66 | <0.001 |
| Sex (male vs. female) | 1.13 | 0.689 | 1.10 | 0.253 |
| BMI at age 15 ($kg/m^2$) | 0.23 | 0.346 | 0.15 | 0.037 |

BP = blood pressure; SBP = systolic BP; DBP = diastolic BP; BMI = body mass index

**Table S6: Stratified models for rate of change in body mass index on systolic and diastolic blood pressure by birth weight category**

| Variable | Low Birth Weight | | Normal Birth Weight | |
|---|---|---|---|---|
| | Regression Coefficient | P-value | Regression Coefficient | P-value |
| **Change in BMI and SBP** | | | | |
| BMI at birth z-score | -4.28 | 0.005 | 0.42 | 0.429 |
| Change in BMI 0-6 months | 1.11 | 0.356 | 0.67 | 0.148 |
| Change in BMI 6 months – 2 years | 2.08 | 0.009 | 1.51 | <0.001 |
| Change in BMI 2 years – 8 years | 1.50 | 0.314 | 1.17 | 0.002 |
| Change in BMI 8 years – 15 years | 1.51 | 0.165 | 0.66 | 0.129 |
| Gestational age at birth (days) | 0.08 | 0.161 | -0.10 | 0.028 |
| Socioeconomic status score at birth | 0.72 | 0.003 | -0.02 | 0.869 |
| Age (years) | -0.45 | 0.404 | -1.11 | <0.001 |
| Sex (male vs. female) | 8.59 | 0.002 | 6.84 | <0.001 |
| Height at age 15 | -0.001 | 0.977 | 0.07 | 0.056 |
| **Change in BMI and DBP** | | | | |
| BMI at birth z-score | -2.63 | 0.013 | 0.05 | 0.888 |
| Change in BMI 0-6 months | -0.71 | 0.334 | 0.49 | 0.116 |
| Change in BMI 6 months – 2 years | 0.42 | 0.535 | 0.65 | 0.044 |
| Change in BMI 2 years – 8 years | 0.33 | 0.761 | 0.64 | 0.025 |
| Change in BMI 8 years – 15 years | 2.06 | 0.022 | 0.57 | 0.102 |
| Gestational age at birth (days) | 0.02 | 0.527 | -0.04 | 0.262 |
| Socioeconomic status score at birth | 0.32 | 0.052 | 0.05 | 0.517 |
| Age (years) | 1.46 | <0.001 | 0.65 | <0.001 |
| Sex (male vs. female) | 1.53 | 0.452 | 0.19 | 0.784 |
| Height at age 15 | -0.02 | 0.525 | 0.02 | 0.489 |

BP = blood pressure; SBP = systolic BP; DBP = diastolic BP; BMI = body mass index

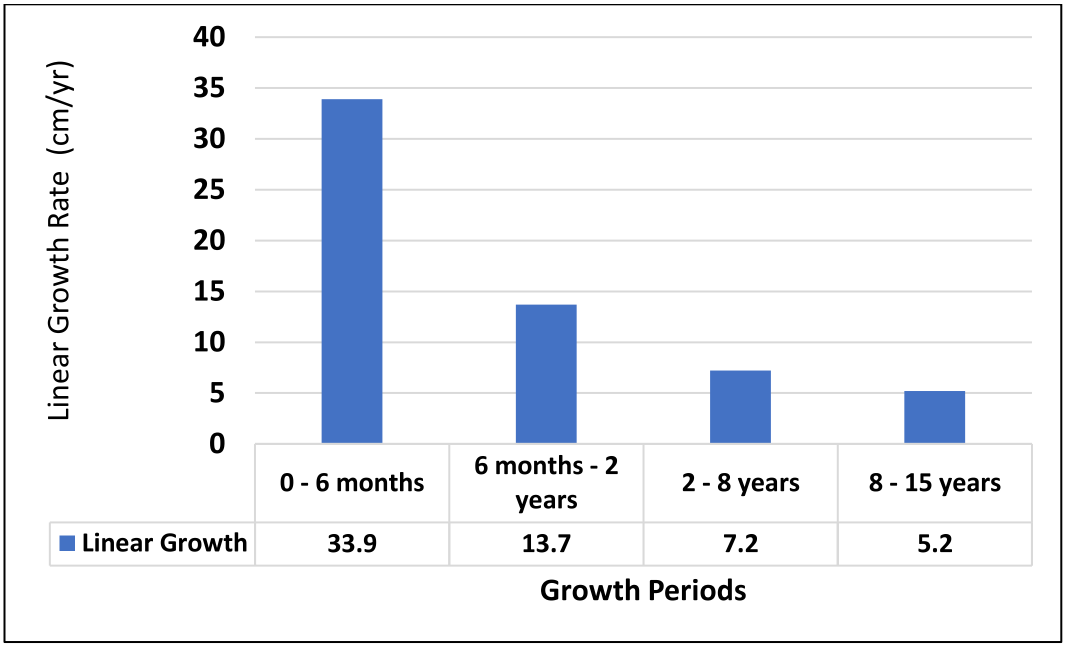

**Figure S1: Annualized mean linear growth rates in age periods during childhood**

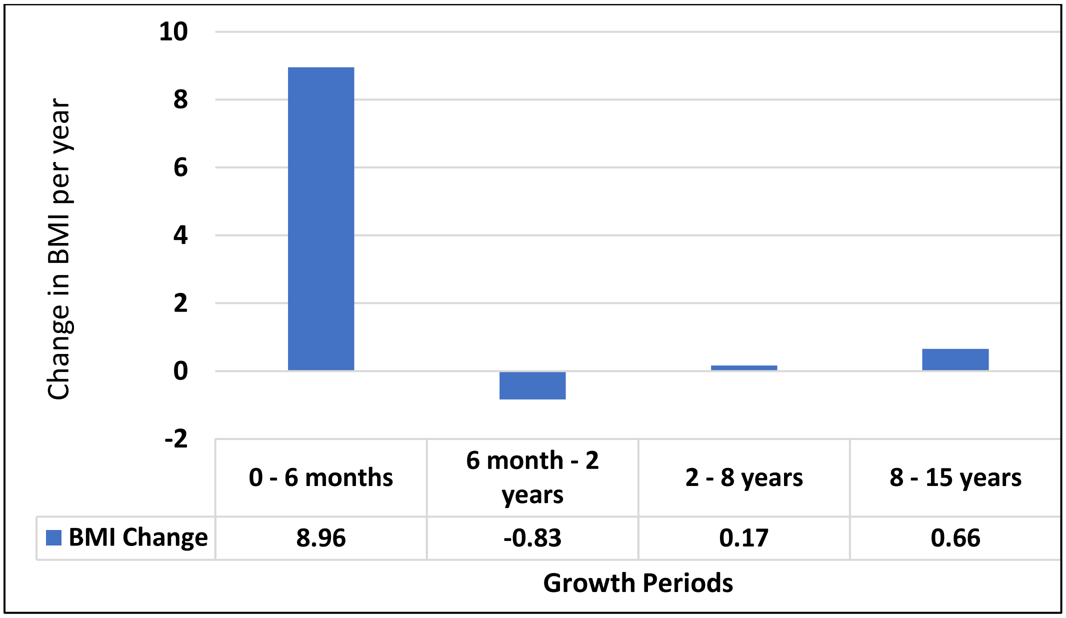

**Figure S2: Mean change in body mass index in age periods during childhood**

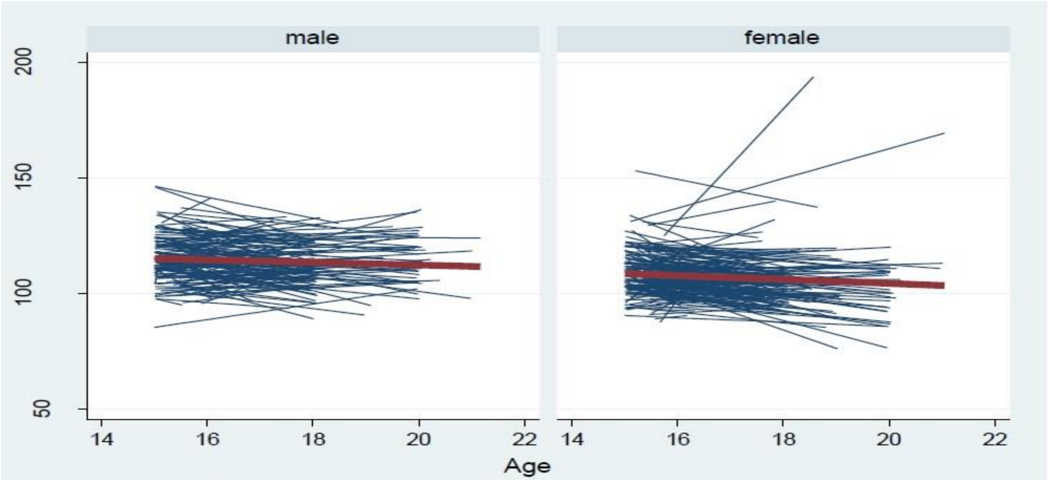

Panel A: Systolic Blood Pressure (mm Hg)

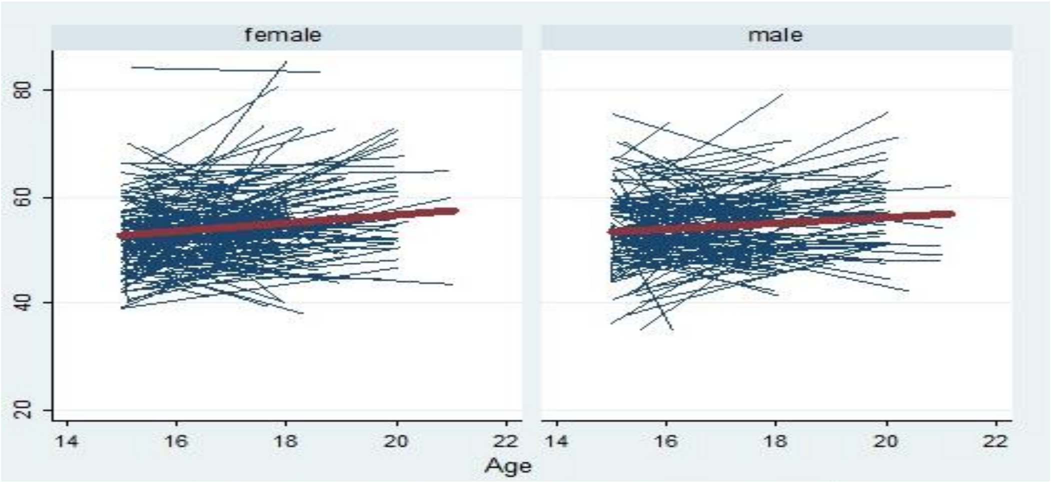

Panel B: Diastolic Blood Pressure (mm Hg)

**Figure S3: Spaghetti plot showing participants' systolic blood pressure (SBP, mm Hg) [Panel A] and diastolic blood pressure (DBP) [Panel B] from age 15-21 years vs age (years) for male and female participants.**

Each line represents the change in the SBP for each participant in the study from 15-21 years. The centre red line represents the mean SBP for all participants together over the period.
